# A reproducible open-source framework for defining type 1 and type 2 diabetes research cohorts in routinely collected electronic health record data

**DOI:** 10.1101/2025.03.26.25324678

**Authors:** Rhian Hopkins, Pedro Cardoso, Laura M Güdemann, Andrew P McGovern, John M Dennis, Beverley M Shields, Katherine G Young

**Affiliations:** Department of Clinical and Biomedical Sciences, University of Exeter Medical School, RILD Building, Royal Devon & Exeter Hospital, Barrack Road, Exeter, EX2 5DW, UK

## Abstract

**Background:** Electronic health record data (EHR) data provide an increasingly important resource for studying people living with diabetes and their clinical outcomes, but robustly coding reproducible datasets is challenging. We aimed to develop a standardised data-processing framework for defining cohorts of people with type 1 and type 2 diabetes using EHR data.

**Methods:** We initially provide a standardised, generalisable procedure to develop clinically reviewed code lists to robustly define variables in EHR data. Using UK population-based data from primary care linked to hospital admission records (Clinical Practice Research Datalink [CPRD]), we develop and demonstrate a data-processing pipeline applicable to raw EHR data, using clinical code lists to define a population of individuals with diabetes and defining their diabetes diagnosis dates using the earliest recorded observation of diabetes (clinical code, high HbA1c test result, or prescription for glucose lowering therapy). Using a previously validated approach, we classify diabetes type (gold standard type 1, type 2) based on insulin prescriptions, diabetes type specific clinical codes, and age at diagnosis. Finally, we demonstrate how multiple research cohorts can be defined from this diabetes population based on a specific index date, including a range of baseline features (sociodemographic and lifestyle factors, biomarkers, comorbidities, medications) and key outcomes relevant to the research question.

**Results:** Application of the framework identified an incident cohort at diabetes diagnosis (type 1 diabetes (T1D): N = 10,480, mean age at diagnosis [SD] = 10.4 [4.8]; type 2 diabetes (T2D): N = 726,800, mean age at diagnosis [SD] = 60.5 [13.4]), a prevalent cohort actively registered with their GP practice on 01/02/2020 (T1D: N = 9,514, T2D: N = 559,905), and a T2D cohort initiating treatment with glucose-lowering therapies (N = 769,394 treatment initiations, considering 7 major medication classes). We publicly share our code lists and data processing code, making our research as transparent and reproducible as possible (https://github.com/Exeter-Diabetes/CPRD-Cohort-scripts, https://github.com/Exeter-Diabetes/CPRD-Codelists/).

**Conclusions:** We have developed a flexible and reproducible framework to generate analysis-ready diabetes research cohorts in EHR data. The concepts of this framework are applicable to any EHR dataset and have been shared for use by other researchers. This approach could improve the quality and reproducibility of the diverse epidemiological and clinical diabetes studies using EHR worldwide.

## Introduction

Diabetes is a major increasing health concern, currently affecting around 830 million people worldwide (1, 2). The condition can result in serious complications such as kidney disease, heart attacks, and amputation (1). These outcomes can often be delayed or prevented through intervention and treatment (1), but the global burden of diabetes is rapidly increasing, and glycaemic control remains suboptimal for large proportions of patients (3–5). This highlights the need for more research.

Electronic health records (EHR) are observational data that are generated and collected as part of routine clinical care, for example from consultation with a GP, or a hospital admission. EHR data are increasingly used in diabetes research and provide an invaluable resource as the data reflects real-world clinical practice. The size of EHR datasets means researchers can define very large cohorts of people with type 1 and type 2 diabetes and allows the study of less common diabetes subtypes. EHR data are representative of the broader population and provide long-term follow-up of patients (6). Therefore, they are ideal for studying clinically relevant outcomes to aid our understanding and management of diabetes. This type of real-world data also enables the study of outcomes and groups of people not covered by clinical trials (7).

Transforming raw EHR data into a dataset ready for analysis requires a large amount of processing and phenotyping, which is complex and time consuming (8). These steps are commonly repeated for each individual study published, even within research groups, with different researchers often using different rules to define the same target population. EHR data processing approaches are usually not standardised, resulting in heterogeneity in cohorts and phenotypes. Full descriptions of data processing are rarely provided in research papers, limiting the reproducibility of their results (8, 9). EHR resources, such as the Clinical Practice Research Datalink (CPRD), often publish data profiles to give researchers information on the structure and contents of the data (10) but guides on how to define specific cohorts are usually not available. As EHR data are becoming increasingly popular in observational diabetes research, it is important to make the data-processing and phenotyping involved as reproducible as possible and to use validated approaches.

We aimed to develop a flexible and reproducible framework for defining standardised type 1 and type 2 diabetes cohorts in EHR data with baseline features and outcomes data. The framework we describe has been informed by over 10 years of experience analysing EHR data as part of the MASTERMIND diabetes precision medicine consortium (11–19).

## Methods

Transforming raw EHR data into a diabetes cohort ready for analysis requires several processing steps. Figure 1 summarises the key steps of our flexible framework to create reproducible diabetes cohorts, which can be adapted and applied to any EHR dataset.

**Figure 1:**
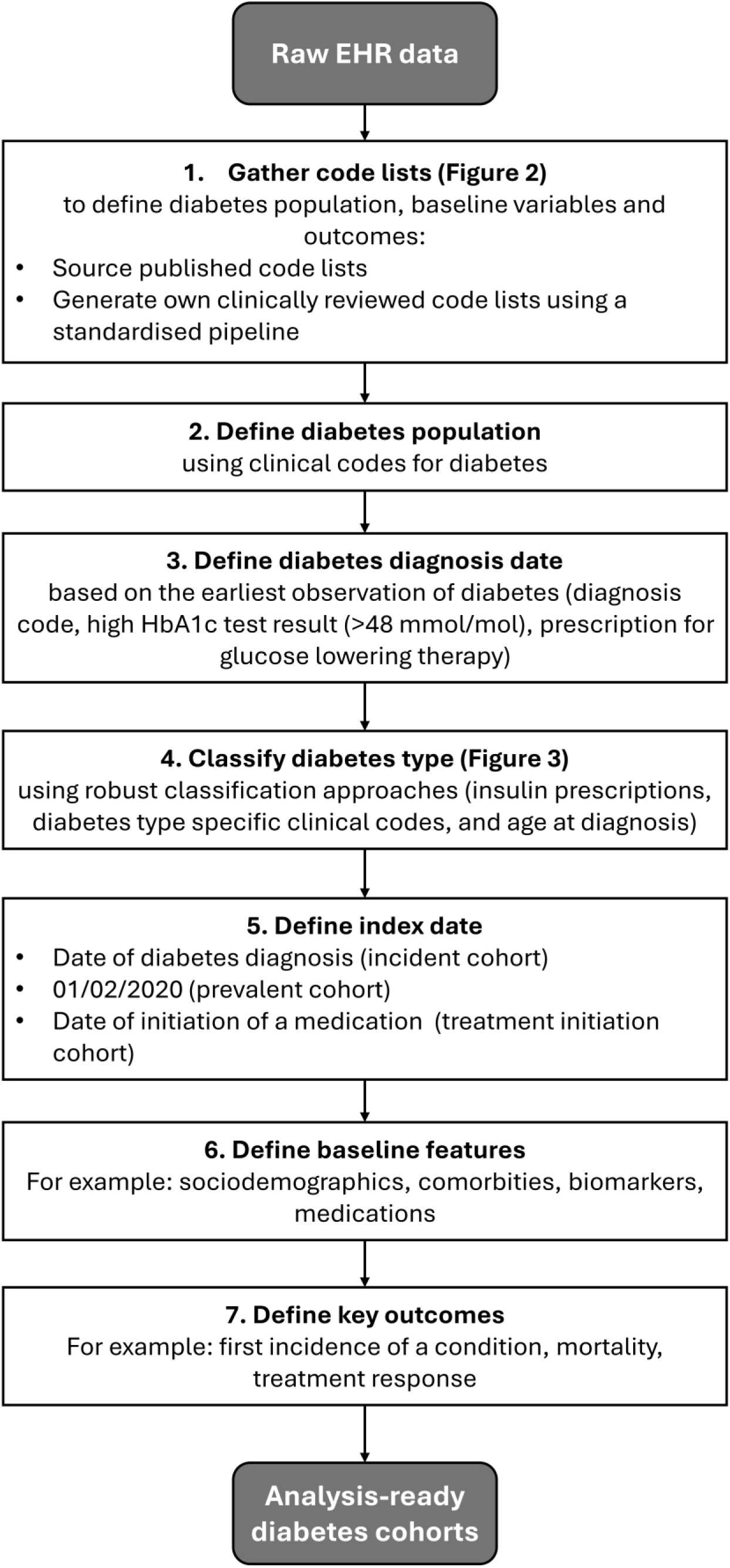
A framework to define diabetes research cohorts using EHR data.

### Data source

We have used EHR data from the Clinical Practice Research Datalink (CPRD) in our diabetes studies. CPRD Aurum is a large database of longitudinal, routinely collected medical records from primary care practices in the UK and contains information on patients’ demographics, diagnoses, prescriptions, lifestyle factors, and test results (10). CPRD Aurum covers over 13% of the UK population and is largely representative of the broader population (10, 20).

We obtained data from CPRD for people who had a record of diabetes between 01/01/2004 and 06/11/2020. The primary care data was linked to other datasets: Hospital Episode Statistics (HES) data which contain information on all admissions to the National Health Service (NHS) secondary care providers, index of multiple deprivation (IMD) data which is the national measure of deprivation, and Office for National Statistics (ONS) Death Registration Data which cover dates and causes of death and is considered the gold standard mortality data in the UK.

### Code lists

EHR data are commonly stored in the form of codes. Therefore, to define each variable of interest in our dataset, we need to generate a list of the codes that could be used to record that variable (for example, a medical condition). These code lists are subsequently used to define the study population, baseline features, and outcomes. While reproducible methods for developing code lists have been suggested (21), standardised approaches are often not used. Published code lists are available for use from online repositories, or they can be generated for the study using a standardised pipeline such as the one we have defined in Figure 2.

**Figure 2:**
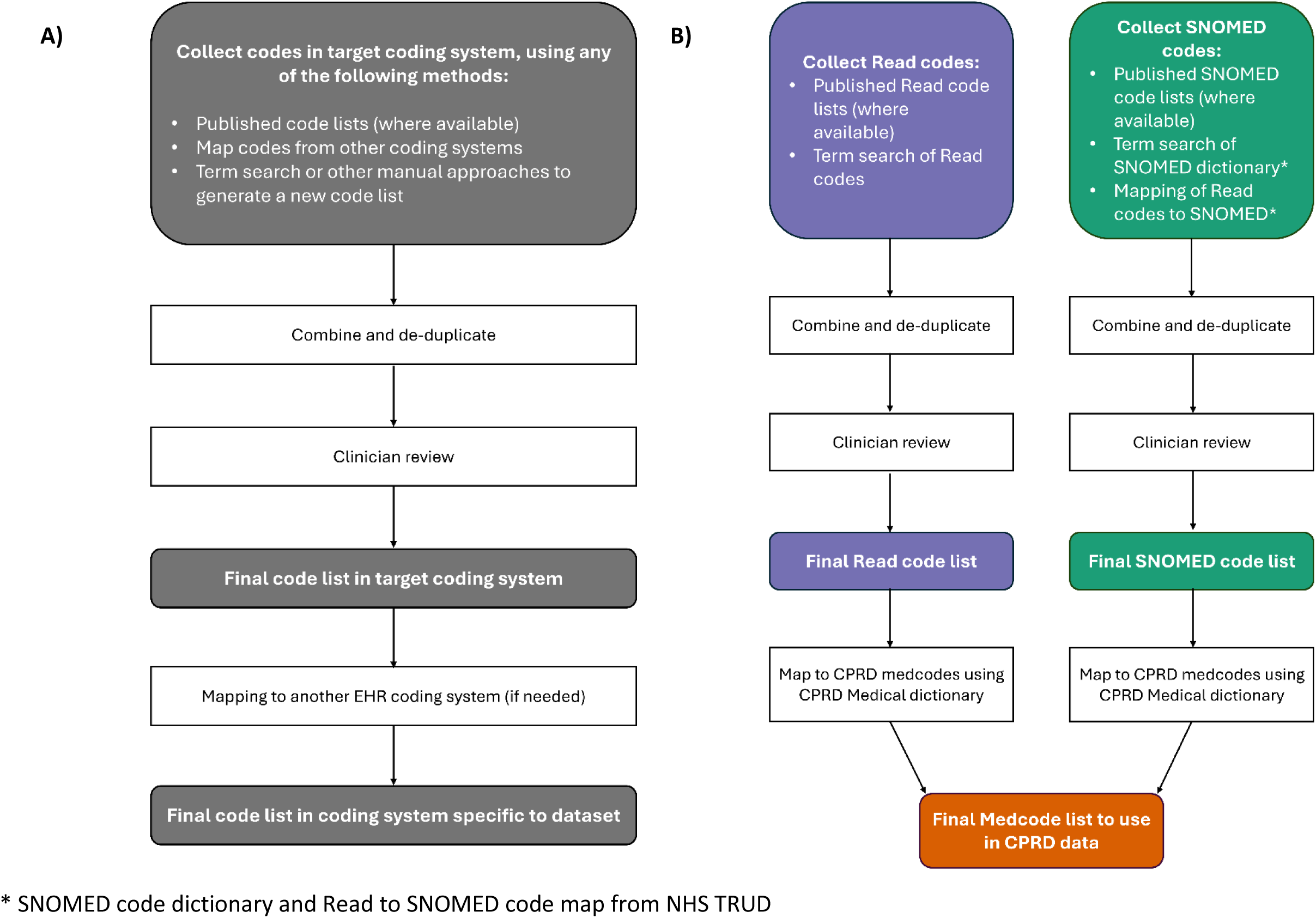
**A) A standardised pipeline to generate clinically reviewed code lists. B) Implementation of pipeline to define Read, SNOMED and CPRD med code lists.**

The pipeline we have developed generates comprehensive code lists that can be used to identify any condition, biomarker, or sociodemographic features. We primarily use existing code lists as inputs, sourced from online repositories (for example, the HDRUK Phenotype library (22) or OpenCodelists (23)), as this improves the efficiency of our approach and reduces research waste. When existing code lists are not available, or to ensure comprehensiveness, term searching the target coding system or mapping from one coding system to another can also provide further codes. Codes from different sources are then combined and reviewed by a clinician. The clinician review is a key step in our process, ensuring our definitions are as robust as possible. This pipeline can easily be applied to any EHR coding system.

We implemented our pipeline using SNOMED codes, the most comprehensive coding system used worldwide (24), and Read codes, historically used in GP systems before switching to SNOMED. Read and SNOMED code lists are initially compiled using existing publicly available code lists where available or term searching the respective coding systems. SNOMED codes are additionally mapped from Read codes and term-search of a SNOMED dictionary to be as comprehensive as possible. As we use CPRD Aurum data for our research, we use CPRD’s Medical Dictionary to map the Read and SNOMED codes to medcodes (CPRD’s own coding format). The generation of Read and SNOMED code lists additionally means this approach is generalisable to other EHR datasets using the same underlying coding system.

To develop code lists for prescriptions, we generated a list of generic and brand names for each medication and searched for these terms in CPRD’s Product Dictionary. A clinician then reviewed the resulting code list.

ICD-10 (for hospital diagnoses in HES and causes of death in ONS) and OPCS-4 (for procedures in HES) code lists were put together using publicly available existing code lists (where available) and adjusted to fit our variable definitions with input from a clinician. All the code lists we have developed are available online in a GitHub repository (https://github.com/Exeter-Diabetes/CPRD-Codelists).

### Defining a diabetes cohort and initial data quality checks

A cohort of people with diabetes can be defined by selecting those with clinical codes for diabetes in their records. NHS England developed the Quality Outcomes Framework (QOF) (25) to provide financial remuneration for good practice for data collection and coding. We included patients who had a diabetes diagnosis code included in the Quality and Outcomes Framework (QOF) list of recommended diabetes codes, recorded with a valid date. This was validated in UK Biobank data(26) using diabetes QOF codes, where our approach had 99.9% specificity and 89.5% sensitivity for self-reported diabetes at nurse interview (personal communication, K Young 2025). Our code lists used to define diabetes in CPRD are available in our GitHub repository: https://github.com/Exeter-Diabetes/CPRD-Codelists/.

CPRD have their own quality standards for data, which include identifying patients with non-continuous follow-up or poor data recording issues (for example: missing year of birth, GP registration date before year of birth, age greater than 115 at end of follow-up) whose records are deemed not ‘acceptable’ for research (27). We therefore excluded these patients from our diabetes cohort.

#### Valid dates

When cleaning the data, we only considered records with a valid date, which we defined as being no earlier than the patient’s date of birth (or month of birth if the date was not available) and no later than the end of the patient’s practice registration, last collection date from the practice, or date of death (when present). If a patient’s full date of birth was not available, we defined their date of birth as the 15^th^ of their month of birth if the birth month and year were available, the 1^st^ July of their birth year if only the birth year was available, or the date of their earliest primary care observation if this was earlier (excluding dates earlier than the month/year of birth).

### Defining diabetes diagnosis dates

Exact diagnosis dates often cannot be directly extracted from EHR data. Therefore, we defined dates of diabetes diagnosis as the earliest observation of diabetes in a person’s record: first diabetes diagnosis code, first HbA1c result of 48 mmol/mol or higher, or first prescription for glucose lowering therapy. If an individual’s diabetes diagnosis date was very close to their GP registration start date (diagnosis within 90 days before or 30 days after registration) we considered this likely unreliable and excluded these individuals from our cohort. We also exclude those with gestational diabetes as calculating their date of diagnosis is more complicated.

### Classifying diabetes type

Classifying diabetes type can be challenging, and in order to distinguish between different types of diabetes in EHR data, robust and validated classification approaches should be used. These can be based on clinical codes, prescriptions, and features such as age at diabetes diagnosis. Figure 3 details a full algorithm of classification approaches that can be used to classify a diabetes type for everyone in a diabetes cohort using EHR data. When classifying type 1 and type 2 diabetes, we defined those with no prescriptions for insulin in their records as type 2. In insulin-treated people, we then evaluated clinical codes specific to a diabetes type. In the majority of cases a person’s diabetes type is clearly defined (over 95% of individuals in our CPRD cohort have codes for solely type 1 diabetes or type 2 diabetes). For the small proportion where a person has clinical codes for multiple types of diabetes, we used the ratio of different type codes to determine diabetes type (based on previously validated studies (28, 29)). Where a patient has no type-specific codes (1.4% of cases), additional approaches may need to be considered, for example age at diabetes diagnosis, time to insulin, and other diabetes medication prescriptions. This approach broadly classifies all individuals by which type of diabetes their clinician likely thought they had. If an individual had a clinical code for another type of diabetes (for example, monogenic diabetes or secondary diabetes) at any date in their records, we classified their diabetes type as ‘other’. Additional work is needed to define the specific diabetes subtype of these individuals (for example, type 3c diabetes (30)).

**Figure 3:**
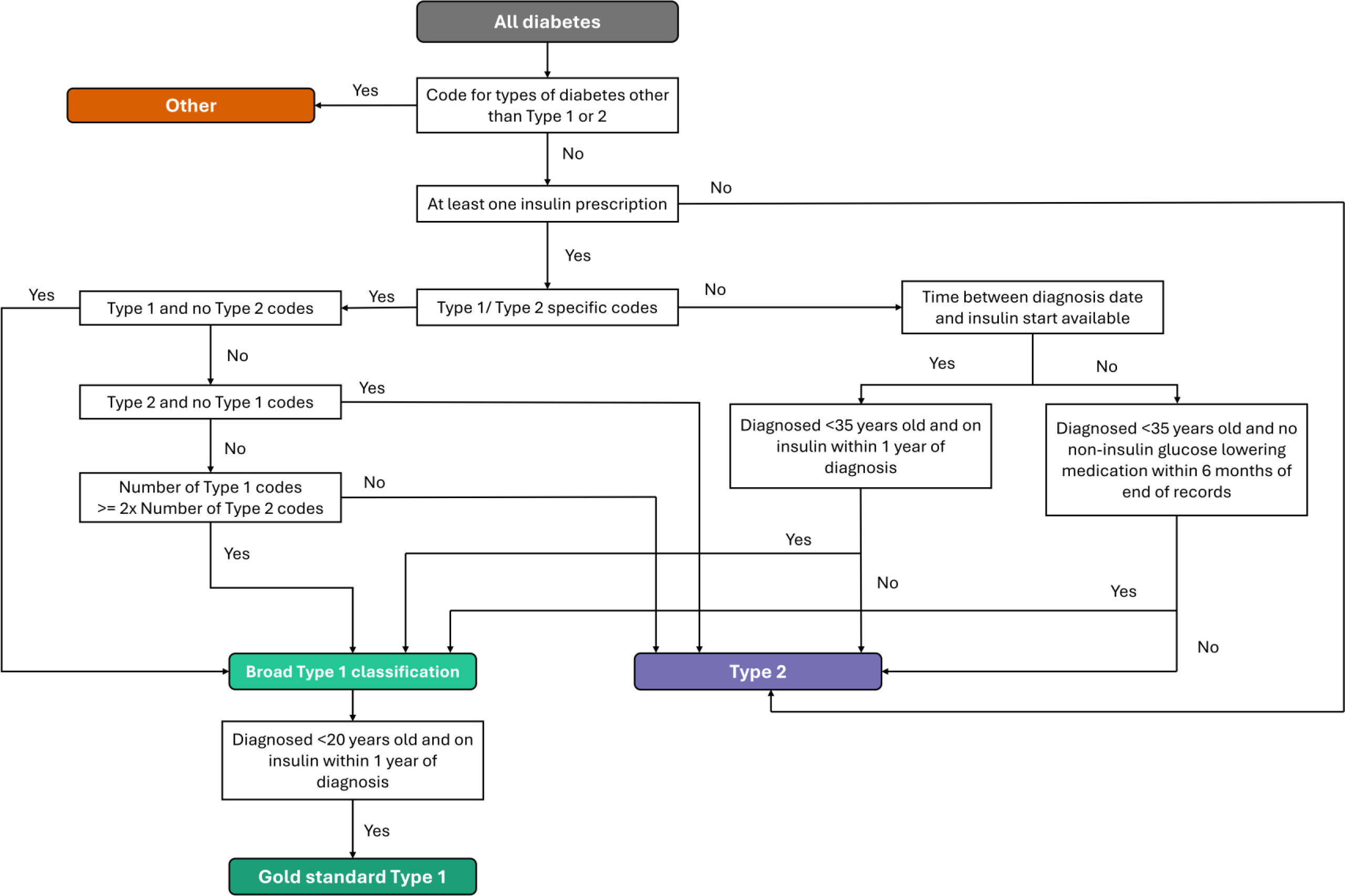
A validated algorithm for classifying type 1 and type 2 diabetes.

In some cases, classifying all patients may not be necessary, and particularly for type 1 diabetes research, it may be more important to define a more specific cohort using stricter criteria.

Therefore, we defined a ‘gold standard’ cohort where we are more confident that they have type 1 diabetes by including only individuals broadly classified as type 1, diagnosed aged 20 or under, and treatment with insulin from within 1 year of diagnosis.

### Defining an index date

Different diabetes cohorts can flexibly be defined using different chosen index dates:

- Incident diabetes cohort: the date of diabetes diagnosis
- Prevalent cohort: A specific time of interest (e.g. 01/02/2020, pre-Covid19)
- Treatment response cohort: The date of initiation of a glucose-lowering medication

### Defining baseline features

Baseline features such as sociodemographics, biomarkers, clinical measurements, and comorbidities can be defined for a cohort relative to the chosen index date. Additional data cleaning and processing steps may be required to define each variable.

#### Biomarkers and clinical measurements

We defined a range of biomarkers for our diabetes cohorts, including: glycated haemoglobin (HbA1c), high-density lipoprotein (HDL), triglycerides, blood creatinine, low-density lipoprotein (LDL), alanine transaminase (ALT), aspartate transferase (AST), total cholesterol, and albumin-creatinine ratio (ACR), as well as the clinical measurements weight, height, body mass index (BMI), systolic blood pressure (SBP), and diastolic blood pressure (DBP). The following data processing steps could also be extended to other biomarkers or clinical measurements. For all biomarkers we removed any values outside of clinically plausible limits. These clinically plausible limits were chosen based on discussion with a clinician and values outside this range were assumed to be errors. We also dropped any observations that did not have acceptable units. For units that made up more than 0.1% of observations, including missing units, we examined the distribution of values to decide if they were acceptable. Where multiple different biomarker values were recorded on the same day, we took the mean of these values. For HbA1c, we additionally removed any observations before 1990 (HbA1c was not widely used before then) and converted all values less than 20 (assumed to be in % units) into mmol/mol. We also calculated estimated glomerular filtration rate (eGFR) from serum creatinine readings using the 2021 CKD-EPI Creatinine Equation (31).

#### Comorbidities

We also defined a range of comorbidities (any condition in addition to diabetes) based on clinical codes, including cardiovascular (hypertension, atrial fibrillation, angina, myocardial infarction, cardiac revascularisation, ischaemic heart disease, heart failure, peripheral vascular disease), respiratory (asthma, chronic obstructive pulmonary disease), neurological (transient ischaemic attack, stroke, dementia, other neurological conditions), oncological (haematological cancer, solid cancer), and other conditions (chronic kidney disease/end-stage renal disease, chronic liver disease, solid organ transplant, rheumatoid arthritis, cystic fibrosis). To define a diagnosis date for a comorbidity we used the earliest date of any record of the condition (in the primary care or hospital data). We also defined microvascular diabetes complications (diabetic nephropathy, neuropathy, retinopathy) using the same method.

#### Sociodemographic and lifestyle factors

We defined ethnicity based on clinical codes in the primary care records and categorised using the same method as Mathur et al in a previous study (32), grouping ethnicity codes into 16 categories that can be collapsed into 5 higher level categories (White, South Asian, Black, Mixed, Other) based on UK census groups. We took the most commonly recorded ethnicity category, and if there was more than one equally most common, we took the most recently recorded. For anyone who we could not define an ethnicity from the primary care records, we used their ethnicity recorded in the hospital data (HES) where it was available. Using this method, we could define an ethnicity for over 97% of our diabetes cohort.

To define deprivation, we used a linked Index of Multiple Deprivation dataset. This is the official measure used in England and assigns each patient a decile representing relative deprivation based on a range of factors (33).

For smoking, we grouped codes into 3 categories (non-smoker, ex-smoker, active smoker) and defined a category for each patient by taking the most recently recorded. If the most recently recorded code is in the ‘non-smoker’ category, but the patient has previously been recorded as a smoker, they are categorised as an ‘ex-smoker’. Similarly for alcohol consumption we categorised codes into 4 groups (none, within recommended limits, excess, harmful) and took the most recently recorded observation.

### Example cohorts and outcomes

Using code lists and a flexible data-processing pipeline as detailed above, we can define various cohorts of people with diabetes for a specific index date, including their baseline features and key outcomes relevant to the research question, for example:

#### Incident cohort

An incident cohort of individuals at the date of diagnosis of their diabetes (diagnosis dates defined as above). We excluded anyone with a diagnosis date before or within 3 months of their date of GP practice registration, reducing the likelihood of including people with an existing diabetes diagnosis transferred to a new GP practice. For this cohort, we can define outcomes such as the first incidence of a comorbidity after diabetes diagnosis, including cardiovascular disease, stroke, and changes in chronic kidney disease stage (in primary care or hospital records), mortality-related outcomes including the date and cause of death defined from the linked ONS data, and time to treatment intensification for example time from diabetes diagnosis to the initiation of insulin treatment.

#### Prevalent cohort (1^st^ February 2020)

A cross-sectional prevalent cohort of individuals with diabetes actively registered with their GP practice on 01/02/2020. We excluded anyone who deregistered or died before 01/02/2020 and anyone whose diabetes was diagnosed after this date. Using the predefined index date, we can then define future outcomes (such as the first incidence of a medical condition, hospitalisation, or mortality).

#### Treatment outcomes

A cohort of all individuals with diabetes initiating a glucose-lowering therapy class at the date of initiation of that medication. For this cohort, we updated the approach previously published by Rodgers *et al*. (34), which detailed the processing steps to define a type 2 treatment cohort using an older version of CPRD data (CPRD GOLD). The drug classes included were metformin, sulphonylureas, thiazolidinediones, DPP4-inhibitors, SGLT2-inhibitors, GLP1-receptor agonists, and insulin. We also extracted acarbose and glinide prescriptions, but due to low numbers prescribed in the UK, these have not been included in our research to date. Individuals could be included more than once if they initiated multiple therapy classes.

We defined a range of short- and long-term treatment outcomes for this cohort. We defined glycaemic response as the change from baseline HbA1c 12 months after drug initiation (the closest measure to 12 months after initiation within 3-15 months) with no addition or cessation of other glucose-lowering medications and continued prescription of the drug of interest. We similarly defined weight change 12 months from initiation on stable unchanged therapy. We also defined early treatment discontinuation as a therapy discontinued within 6 months of initiation. As described by Rodgers *et al*., gaps between prescriptions had to be at least 6 months to be considered as a drug being stopped, and the availability of at least 3 months follow-up time after discontinuation was required to confirm the drug was discontinued (34). We additionally defined glycaemic failure (confirmed HbA1c ≥69 mmol/mol), time to treatment change, and time to insulin initiation. We also defined a range of long-term outcomes, including all-cause mortality, major cardiovascular events or heart failure, renal progression (>40% decrease in eGFR or end-stage kidney disease), and microvascular complications.

Scripts to define these example cohorts and generic template scripts that can be adapted to define other cohorts are available online in our GitHub repository (https://github.com/Exeter-Diabetes/CPRD-Cohort-scripts).

## Results

From CPRD, we obtained an October 2020 download including primary care data on 1,480,985 individuals with a record of diabetes between 01/01/2004 and 06/11/2020 (1,138,193 after applying quality control measures), and any linked hospital, mortality and deprivation data where available. Figure 4 describes the inclusion criteria and resulting numbers in our diabetes dataset and three example cohorts.

**Figure 4:**
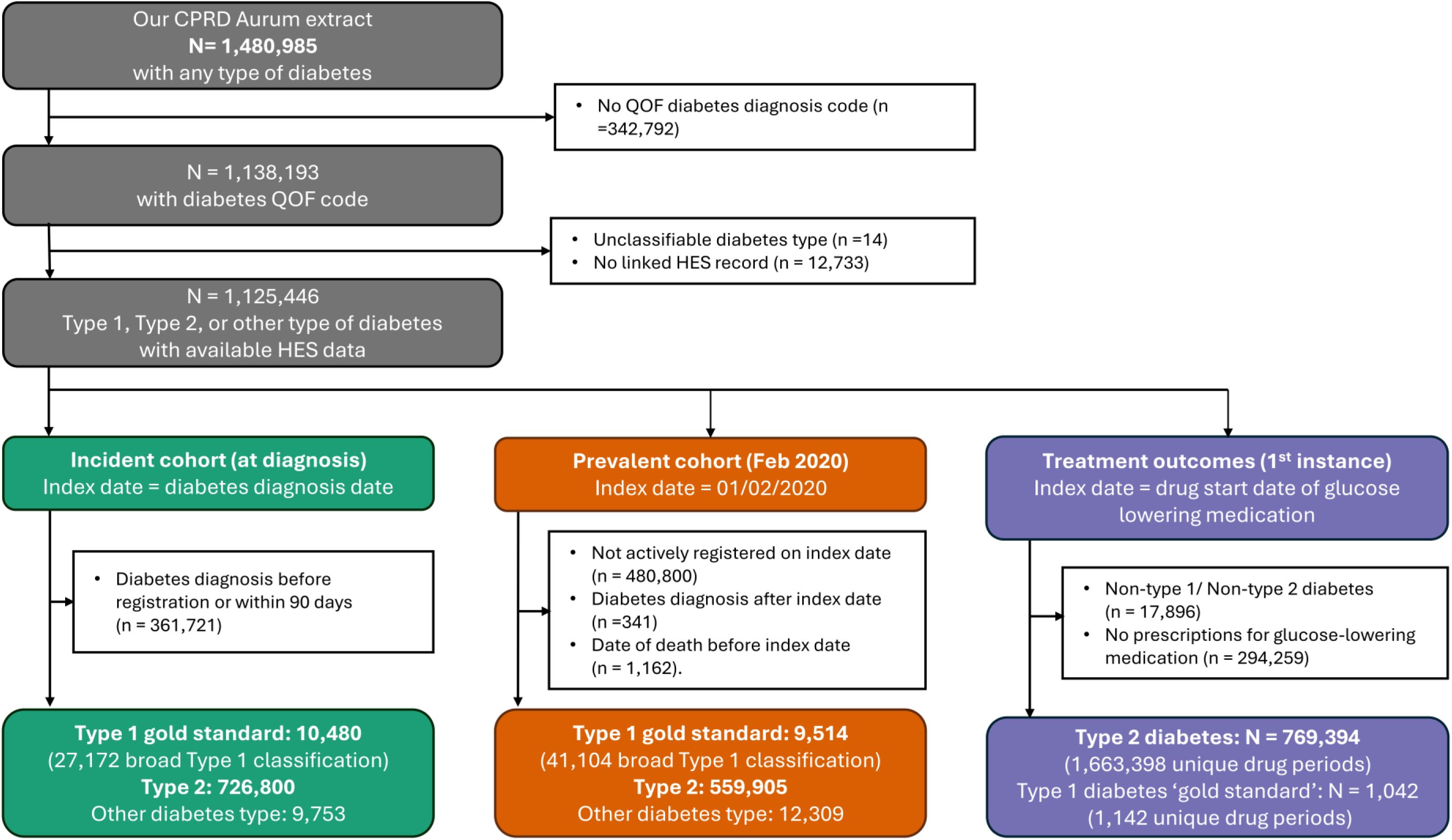
**Diabetes research cohorts (incident cohort, prevalent cohort, and treatment outcomes cohort) derived from CPRD primary care records and linked datasets.**

### Cohorts

The incident cohort, with data covering diabetes diagnosis, consisted of 10,480 people with type 1 diabetes ‘gold standard’, 726,800 with type 2 diabetes, and 9,753 with other types of diabetes. Table 1 describes the key baseline characteristics of this cohort.

**Table 1:** Baseline characteristics of an incident cohort of gold standard people with type 1 diabetes and type 2 diabetes at diagnosis.

|  | **T1 gold standard** | **T2** |
| --- | --- | --- |
| n | 10,480 | 726,800 |
| **Follow up, years** |  |  |
| Median [IQR] | 7.9 [4.1, 13.3] | 8.4 [4.6, 13.7] |
| **Sex** |  |  |
| Male | 5,818 (55.5) | 405,683 (55.8) |
| Female | 4,662 (44.5) | 321,117 (44.2) |
| **Age, years** |  |  |
| Mean (SD) | 10.4 (4.8) | 60.5 (13.4) |
| **Ethnicity** |  |  |
| White | 9,099 (86.8) | 595,521 (81.9) |
| South Asian | 502 ( 4.8) | 66,510 ( 9.2) |
| Black | 444 ( 4.2) | 32,687 ( 4.5) |
| Other | 141 ( 1.3) | 8,703 ( 1.2) |
| Mixed | 192 ( 1.8) | 5,822 ( 0.8) |
| Unknown | 102 ( 1.0) | 17,557 ( 2.4) |
| **Deprivation quintile** |  |  |
| 1 | 2,207 (21.1) | 131,204 (18.1) |
| 2 | 2,003 (19.1) | 137,245 (18.9) |
| 3 | 1,905 (18.2) | 140,728 (19.4) |
| 4 | 2,118 (20.2) | 153,350 (21.1) |
| 5 | 2,243 (21.4) | 163,724 (22.5) |
| Missing | 4 ( 0.0) | 549 ( 0.1) |
| **HbA1c, mmol/mol** |  |  |
| Mean (SD) | 104.6 (29.7) | 62.4 (22.5) |
| **BMI, kg/m2** |  |  |
| Mean (SD) | 21.3 (6.2) | 32.3 (6.8) |

The prevalent cohort, actively registered with their GP practice on 01/02/2020, consisted of 9,514 people with type 1 diabetes ‘gold standard’, 559,905 with type 2 diabetes, and 12,309 with other types of diabetes). Table 2 describes the key baseline characteristics of this cohort.

**Table 2:** Baseline characteristics of a prevalent (2020) cohort of people with gold standard type 1 diabetes and type 2 diabetes.

|  | **T1 gold standard** | **T2** |
| --- | --- | --- |
| n | 9,017 | 559,905 |
| **Sex** |  |  |
| Male | 5,016 ( 55.6) | 316,232 (56.5) |
| Female | 4,001 ( 44.4) | 243,673 (43.5) |
| **Age, years** |  |  |
| Mean (SD) | 20.5 (8.4) | 67.4 (13.1) |
| **Duration of diabetes, years** |  |  |
| Mean (SD) | 10.1 (7.0) | 10.8 (7.5) |
| **Ethnicity** |  |  |
| White | 7,859 ( 87.2) | 429,884 (76.8) |
| South Asian | 430 ( 4.8) | 72,099 (12.9) |
| Black | 376 ( 4.2) | 32,414 ( 5.8) |
| Other | 119 ( 1.3) | 9,069 ( 1.6) |
| Mixed | 170 ( 1.9) | 5,809 ( 1.0) |
| Unknown | 63 ( 0.7) | 10,630 ( 1.9) |
| **Deprivation quintile** |  |  |
| 1 | 1,991 ( 22.1) | 99,731 (17.8) |
| 2 | 1,751 ( 19.4) | 103,030 (18.4) |
| 3 | 1,611 ( 17.9) | 108,778 (19.4) |
| 4 | 1,776 ( 19.7) | 120,819 (21.6) |
| 5 | 1,888 ( 20.9) | 127,270 (22.7) |
| Missing | NA | 277 ( 0.0) |
| **HbA1c, mmol/mol** |  |  |
| Mean (SD) | 70.5 (19.3) | 57.8 (16.4) |
| **BMI, kg/m2** |  |  |
| Mean (SD) | 24.4 (5.7) | 31.0 (6.6) |
| **Diabetes medications** |  |  |
| Insulin | 9,017 (100.0) | 70,047 (12.5) |
| Metformin | 269 ( 3.0) | 343,592 (61.4) |
| Sulphonyurea | 4 ( 0.0) | 97,546 (17.4) |
| TZD | NA | 10,441 ( 1.9) |
| DPP4-inhibitor | 2 ( 0.0) | 94,051 (16.8) |
| SGLT2-inhibitor | 21 ( 0.2) | 52,763 ( 9.4) |
| GLP-1 agonist | 18 ( 0.2) | 24,568 ( 4.4) |

The type 2 diabetes treatment response cohort consisted of 769,394 people initiating a class of glucose-lowering medication: metformin (n= 636,001), sulphonylureas (n= 352,671), thiazolidinediones (n= 109,249), DPP4-inhibitors (n= 219,755), GLP1-receptor agonists (n= 60,358), SGLT2-inhibitors (n= 99,994), and insulin (n= 162,187). Table 3 describes key baseline characteristics of individuals with type 2 diabetes at initiation of a glucose-lowering therapy by drug class. There were also 1,042 people with type 1 diabetes ‘gold standard’ initiating a class of non-insulin glucose-lowering medication (all concurrently insulin-treated): metformin (n= 985), sulphonylureas (n= 16), thiazolidinediones (n= 7), DPP4-inhibitors (n= 21), GLP1-receptor agonists (n= 54), SGLT2-inhibitors (n= 59).

**Table 3:** Baseline characteristics for a type2 diabetes treatment outcomes cohort at drug initiation, by major drug classes.

|  | Metformin | Sulphonylurea | TZD | DPP4i | SGLT2i | GLP1 | Insulin |
| --- | --- | --- | --- | --- | --- | --- | --- |
| n | 636,001 | 352,671 | 109,249 | 219,755 | 99,994 | 60,358 | 162,187 |
| **Sex** |  |  |  |  |  |  |  |
| Male | 361,409<br>(56.8) | 201,855 (57.2) | 62,562<br>(57.3) | 127,171<br>(57.9) | 60,174<br>(60.2) | 32,231<br>(53.4) | 89,844<br>(55.4) |
| Female | 274,592<br>(43.2) | 150,816 (42.8) | 46,687<br>(42.7) | 92,584<br>(42.1) | 39,820<br>(39.8) | 28,127<br>(46.6) | 72,343<br>(44.6) |
| **Age, years** |  |  |  |  |  |  |  |
| Mean (SD) | 60.9 (13.2) | 61.8 (13.0) | 61.6 (12.2) | 64.0 (13.0) | 58.5 (10.7) | 57.7 (11.1) | 62.8 (13.9) |
| **Ethnicity** |  |  |  |  |  |  |  |
| White | 506,740<br>(79.7) | 284,165 (80.6) | 87,304<br>(79.9) | 168,978<br>(76.9) | 76,350<br>(76.4) | 51,435<br>(85.2) | 132,078<br>(81.4) |
| South Asian | 68,562<br>(10.8) | 36,325 (10.3) | 12,924<br>(11.8) | 29,754<br>(13.5) | 14,399<br>(14.4) | 4,839 ( 8.0) | 15,256 ( 9.4) |
| Black | 32,476 ( 5.1) | 18,160 ( 5.1) | 4,984 ( 4.6) | 12,669 ( 5.8) | 4,714 ( 4.7) | 2,350 ( 3.9) | 9,687 ( 6.0) |
| Other | 8,359 ( 1.3) | 4,100 ( 1.2) | 1,111 ( 1.0) | 3,053 ( 1.4) | 1,532 ( 1.5) | 540 ( 0.9) | 1,754 ( 1.1) |
| Mixed | 5,681 ( 0.9) | 2,975 ( 0.8) | 895 ( 0.8) | 2,120 ( 1.0) | 1,036 ( 1.0) | 517 ( 0.9) | 1,484 ( 0.9) |
| Unknown | 14,183 ( 2.2) | 6,946 ( 2.0) | 2,031 ( 1.9) | 3,181 ( 1.4) | 1,963 ( 2.0) | 677 ( 1.1) | 1,928 ( 1.2) |
| **Deprivation quintile** |  |  |  |  |  |  |  |
| 1 | 109,061<br>(17.1) | 59,646 (16.9) | 18,053<br>(16.5) | 35,781<br>(16.3) | 16,609<br>(16.6) | 9,599<br>(15.9) | 26,288<br>(16.2) |
| 2 | 116,319<br>(18.3) | 64,294 (18.2) | 19,897<br>(18.2) | 38,930<br>(17.7) | 17,466<br>(17.5) | 10,480<br>(17.4) | 28,772<br>(17.7) |
| 3 | 122,473<br>(19.3) | 68,024 (19.3) | 21,261<br>(19.5) | 42,013<br>(19.1) | 19,106<br>(19.1) | 11,473<br>(19.0) | 30,726<br>(18.9) |
| 4 | 137,775<br>(21.7) | 77,119 (21.9) | 23,758<br>(21.7) | 49,142<br>(22.4) | 22,420<br>(22.4) | 13,276<br>(22.0) | 35,634<br>(22.0) |
| 5 | 149,903<br>(23.6) | 83,319 (23.6) | 26,203<br>(24.0) | 53,729<br>(24.4) | 24,332<br>(24.3) | 15,486<br>(25.7) | 40,647<br>(25.1) |
| Missing | 470 ( 0.1) | 269 ( 0.1) | 77 ( 0.1) | 160 ( 0.1) | 61 ( 0.1) | 44 ( 0.1) | 120 ( 0.1) |
| **HbA1c, mmol/mol** |  |  |  |  |  |  |  |
| Mean (SD) | 70.1 (21.0) | 76.5 (21.8) | 74.3 (16.9) | 72.7 (17.2) | 77.6 (17.2) | 78.3 (17.9) | 85.6 (23.6) |
| **BMI, kg/m2** |  |  |  |  |  |  |  |
| Mean (SD) | 32.1 (6.8) | 31.0 (6.6) | 31.4 (6.5) | 31.9 (6.7) | 33.8 (6.9) | 37.6 (7.2) | 30.7 (6.8) |

### Open research

We have shared our R code for defining different diabetes cohorts and our code lists online in GitHub repositories (https://github.com/Exeter-Diabetes/CPRD-Cohort-scripts, https://github.com/Exeter-Diabetes/CPRD-Codelists/), alongside descriptive documentation to help others to understand and replicate our work. Making our work publicly available means our research is as transparent and reproducible as possible, adhering to the principles of open research and FAIR data (35).

## Discussion

We have developed a reproducible framework for defining standardised cohorts of people with type 1 and type 2 diabetes in coded EHR data. The data-processing pipeline can flexibly generate diabetes cohorts from raw datasets that include information about a person’s diabetes, as well as sociodemographic features, comorbidities, biomarkers, and medications. This framework has been implemented in the definition of cohorts for analysis in several published studies (30, 36–40), mainly focussed on glucose-lowering therapy in type 2 diabetes, and has ensured our work is highly reusable and coding is robust and consistent across our studies. EHR datasets are a greatly valuable research resource internationally, and the wide application of our open-source framework could enhance the quality and reproducibility of diabetes epidemiological and clinical studies.

Research using EHR data is becoming increasingly popular, and data sources are becoming more readily available, both in the UK through initiatives such as NHS England’s Secure Data Environment (41) and internationally. Therefore, promoting open research practices and ensuring research is reproducible using standardised data-processing approaches will be increasingly important. While the framework we have outlined focuses on diabetes, the general approaches could be applied to other medical conditions and areas of health research.

## Limitations

EHR data are not collected for research purposes. Therefore, robustly defining variables can be difficult (42), and misclassification is often possible. While our approaches will not be perfect, we have aimed to make them as robust as possible given the constraints of the data, developing thorough algorithms for defining key variables through discussion with clinicians involved in this research. Limitations of using EHR data for research include data completeness and missing data, the reliance on the accuracy of the data recorded in clinical records, and variation in coding between practices over time (6). Exact dates of diabetes diagnosis are not available in CPRD primary care data. We used a combination of diabetes diagnosis codes, HbA1c test results, and prescriptions for glucose lowering therapies to define a diagnosis date as comprehensively as possible, but there still may be cases where this does not accurately reflect the initial diagnosis date. Similarly, an individual’s diabetes type can be difficult to determine in healthcare records (7), where classification biomarker tests (for example, C-peptide) are not commonly available. To deal with this, we used an approach based on previous studies which were validated against clinical assessment (28, 29). We also included both a broad definition of type 1 diabetes that likely reflects a clinical diagnosis and further criteria to define a more restricted cohort where we are more confident that these individuals have type 1. Additionally, determining whether an individual has certain comorbidities can be challenging and relies on the clinical codes used to define the conditions (8). We used comprehensive clinically reviewed code lists (utilising both the primary care data and hospital records) to define each medical condition as robustly as possible. When defining medications, we may miss some prescriptions issued in a hospital or private healthcare, however prescription data are generally well coded in UK primary care records. These highlighted problems may limit the accuracy of some coding but are likely to lead to noise rather than biases in the data, which will be mitigated by the large sample sizes available.

## Conclusion

EHR data can be a highly valuable resource for studying diabetes, but robustly coding reproducible datasets is challenging. Therefore, we developed a flexible and reproducible framework to generate standardised diabetes cohorts using EHR data that we have made publicly available for use worldwide. Whilst we have used UK EHR data from CPRD in this study, our framework is applicable to any coded EHR dataset.

## Acknowledgements

This article is based on data from the Clinical Practice Research Datalink obtained under licence from the UK Medicines and Healthcare products Regulatory Agency. CPRD data are provided by patients and collected by the NHS as part of their care and support. Approval for CPRD data access and the study protocol was granted by the CPRD Independent Scientific Advisory Committee (eRAP protocol number: 22_002000). This study was supported by the National Institute for Health and Care Research Exeter Biomedical Research Centre. The views expressed are those of the authors and not necessarily those of the NHS, NIHR or the Department of Health and Social Care.

## Author contributions

The study concept and design were developed by KGY, BMS, APM, JMD, and RH. KGY and RH prepared and process the data and defined the study cohorts. All authors critically revised the manuscript and saw and approved the final article. KGY and BMS attest that all listed authors meet authorship criteria, that no others meeting the criteria have been omitted. KGY and BMS are responsible for the decision to submit for publication, and are the guarantors of this work and, as such, had full access to all the data in the study and takes responsibility for the integrity of the data and the accuracy of the data analysis.

## Ethics/ data approval

The study protocol was approved by the CPRD Independent Scientific Advisory Committee (eRAP protocol numbers: 22_002000).

## Data availability

No additional data are available from the authors although CPRD data are available by application to CPRD Independent Scientific Advisory Committee.

## Rights Retention

For the purpose of open access, the author has applied a Creative Commons Attribution (CC BY) licence to any Author Accepted Manuscript version arising from this submission.

